# Phenyllactic Acid is Physiologically Released from Skeletal Muscle and Contributes to the Beneficial Effects of Physical Exercise in Humans

**DOI:** 10.1101/2024.03.29.24305064

**Authors:** Miriam Hoene, Xinjie Zhao, Chunxiu Hu, Andreas L. Birkenfeld, Andreas Peter, Andreas M. Nieß, Anja Moller, Qi Li, Rainer Lehmann, Peter Plomgaard, Guowang Xu, Cora Weigert

## Abstract

**Aims/hypothesis:** While physical activity is clearly beneficial in combating type 2 diabetes, the underlying molecular mechanisms are incompletely understood. Moreover, there is a considerable degree of variability in the individual response to exercise-based lifestyle interventions that remains to be explained. We aimed to identify novel exercise-induced metabolites that could mediate the improvement in glycemic control and reduction of obesity and contribute to individual differences in the response to exercise interventions.

**Methods:** We studied acute exercise- and training-induced changes in plasma metabolites in sedentary subjects with overweight (8 male, 14 female) participating in an eight-week supervised training program flanked by two acute endurance exercise sessions. Plasma metabolites were quantified using LC- and CE-MS. In a separate study (n=9 lean males), we assessed metabolite fluxes over the leg using arterial and venous catheters. Functional analyses were performed in primary blood mononuclear cells (PBMCs) stimulated with lipopolysaccharide (LPS) or the saturated fatty acid palmitate.

**Results:** The amino acid breakdown products 3-phenyllactic acid (PLA), 4-hydroxyphenyllactic acid and indolelactic acid were increased after both acute exercise and training. All three aromatic lactic acids, which so far mainly received attention as bacterial metabolites, exhibited an efflux from the leg. PLA showed the largest increase after both acute exercise and training, of 57% and 20% respectively. The magnitude of the acute exercise-induced increase in PLA correlated with a decrease in subcutaneous adipose tissue volume and an improvement in insulin sensitivity over the course of the intervention. Furthermore, both isomers, D- and L-PLA, counteracted inflammatory cytokine production in PBMCs.

**Conclusions/interpretation:** Our findings indicate that PLA is physiologically released from skeletal muscle and can contribute to the anti-inflammatory effects of exercise as well as to individual difference in the response to lifestyle interventions in humans. PLA and potentially, aromatic lactic acids in general may be particularly relevant metabolic regulators because they can be produced both endogenously and by the microbiome.

**Trial registration:** ClinicalTrials.gov <u>NCT03151590</u>

## Introduction

The positive effects of physical activity in improving impaired glucose tolerance and insulin resistance, and thus type 2 diabetes and related cardiometabolic diseases, are closely linked to a reduction in body fat and an improvement of the inflammation milieu [1]. However, the molecular mechanisms underlying the beneficial responses to exercise are still incompletely understood. Moreover, there is a large variability in individual responses to exercise-based lifestyle interventions in terms of therapeutic goals, such as improved glycaemic control or reduction of adipose tissue mass, which has not yet been satisfactorily explained [2–5].

In addition to local and neurohormonal mechanisms that mediate the acute and long-term responses to physical activity, cytokines [6] and more lately, metabolites have been found to play a role as exercise signals [7–9]. These molecules exert para- and endocrine effects when released by contracting skeletal muscle and other organs. Therefore, exercise-induced metabolites may also contribute to differences in the individual response to exercise interventions. Recently, we found post-exercise levels of N-lactoyl-phenylalanine (Lac-Phe), a pseudo-dipeptide shown to lower body weight in mice [10], to correlate with adipose tissue loss in subjects with obesity [11]. Other examples of exercise-regulated intermediary metabolites with extracellular signalling function include succinate, lactate, and beta-aminoisobutyric acid [8, 12–14]. In addition to endogenous metabolites, small molecules produced by the microbiota have been shown to modulate host energy metabolism and furthermore, immune cell function and inflammation [15, 16].

The relevance of exercise-induced factors as metabolic regulators may be particularly high if they are increased not only after acute exercise, but remain at a higher level in trained subjects at rest. To identify such metabolites, we analysed plasma samples obtained in the resting and acutely exercised states in a cohort of subjects with overweight and obesity before and after eight weeks of supervised endurance training. Metabolomic profiling was performed using capillary electrophoresis-time-of-flight mass spectrometry (CE-TOF/MS) and ultra-high performance chromatography-quadruple-time-of-flight mass spectrometry (UHPLC-QTOF/MS). This was combined with functional assays in primary blood mononuclear cells (PBMCs) from healthy donors and with a study in catheterized subjects designed to assess whether the exercise-induced metabolites are released from skeletal muscle.

## Methods

### Exercise intervention study

Healthy, sedentary subjects (less than 120 minutes physical activity per week) with a BMI of 31.7 ± 4.5 kg/m^2^ (mean ± SD; range 27.5–45.5 kg/m^2^) (<u>electronic supplementary material (ESM)</u> Table 1) were recruited for an eight-week supervised exercise intervention. Written consent was obtained from all participants and the study was approved by the ethics committee of the University of Tübingen and registered at Clinicaltrials.gov (NCT03151590).

Details of study, enrolment [17] and sample availability for metabolomics (n=22) have been published recently [11]. Briefly, the intervention consisted of three times per week one hour endurance exercise at 80% VO_2_peak (30 min each cycling and walking). Before and 5 days after the last exercise session, insulin sensitivity was assessed using an 75 g-oral glucose test by the method of Matsuda and DeFronzo (ISI_Mats_), and adipose tissue volume using magnetic resonance imaging (MRI), as described in the <u>ESM</u> Table 1 [11, 17]. Subjects were instructed not to change their dietary habits for the duration of the study. Plasma samples for metabolomics analyses were collected at two acute exercise visits, one at the beginning and one at the end of the eight-week training intervention. At each acute exercise visit, a first blood sample was obtained in the morning in the fasting state and a second one immediately after 30 min of bicycle ergometer exercise at 80% VO_2_peak. Between resting blood sampling and commencement of exercise, the subjects received a standardized breakfast consisting of a bun with butter and cheese, 150 g apple purée, and water.

### Exercise study with catheterization of femoral veins and artery

Arterio-venous metabolite differences over the legs were assessed in a separate study in healthy male, recreationally active subjects (n=9, 20.9 ± 0.5 years, BMI, 22.6 ± 0.8 kg/m^2^) that has been described in detail previously [18]. The study was executed in accordance with the Helsinki Declaration and were approved by the Scientific Ethics Committee of the capital region of Denmark. Briefly, catheters were inserted retrogradely into the femoral artery of one and the femoral veins of both legs and plasma samples collected before, during and after two hours of one-legged kicking exercise. Metabolite fluxes were calculated as described using the mean of blood flow measurements performed in three of the subjects [18]. The study was performed in the morning and subjects remained in the fasting state throughout the test.

### Analysis of plasma metabolites

Metabolite profiling was performed using CE-TOF/MS and UHPLC-TOF/MS. Data acquisition of plasma samples was carried out in both cation-positive and anion-negative mode for both CE- and LC-MS. Details regarding extraction of EDTA plasma samples and metabolite profiling are provided in the <u>ESM Methods</u>.

Metabolites were identified using either standards added before sample extraction, or a CE-MS database containing 960 metabolite standards provided by Human Metabolome Technologies, Inc. (HMT), or an in-house LC-MS database with retention time, MS1 and MS2, as detailed in the <u>ESM</u> Table 2.

### Isolation of leukocyte populations, functional studies in PBMCs and cytokine analyses

EDTA blood was obtained from healthy individuals recruited among the staff of the University Hospital Tübingen. Exclusion criteria were acute or chronic diseases including acute allergic reactions. All volunteers provided written consent to participate in the study. The study was approved by the ethics committee of the University Hospital Tübingen. PBMCs were obtained by centrifugation of PBS-diluted blood on Ficoll–Paque (171440, Cytiva) and washed twice in PBS. Cells were resuspended at a density of 2 Mio cells/ml in RPMI 1640 medium (R0883, Thermo Fisher) supplemented with 2 mM L-Glutamine (G7513, Sigma) and 10% FBS (S0615, Sigma) and incubated at 37 °C, 5% CO_2_ in a humidified atmosphere. After pre-treatment with D-PLA (376906, Sigma) or L-PLA (113069, Sigma) (10-100 µM, as indicated) for 1 h, PBMCs were additionally stimulated with 100 ng/mL lipopolysaccharide (LPS) from *Escherichia coli* (L8274, Sigma) or 500 µM palmitic acid (P0500, Sigma, stock solution 6 mM in FBS) for 2 or 18 h. After incubation, adherent and potential non-adherent cells were combined for RNA isolation and cleared supernatants stored at -80 °C for analysis of released cytokines.

Neutrophils were isolated using Mono-Poly-Medium (ICN, 1698049) as previously described [19].

RNA from PBMCs and neutrophils was isolated using the RNeasy mini kit (Qiagen) according to the manufacturer’s instructions. cDNA was transcribed using the Transcriptor First Strand cDNA Synthesis Kit (Roche). Real-time qPCR was carried out on a LightCycler 480 with QuantiFast reagent (20405, Qiagen) and QuantiTect Primer Assays (*IL1b*: QT00021385, *IL6*: QT00083720, *TNF*: QT01079561, all Qiagen). Standards for each primer were generated by purifying PCR products with the MiniElute PCR Purification Kit (Qiagen) and mRNA levels quantified using a standard curve with known concentrations. mRNA levels were normalized to the reference transcript RPS28 and standardized to the maximal value per donor before calculating mean values for technical replicates (n = 2 per donor and treatment).

IL-6 was measured on the ADVIA Centaur XPT immunoassay system (Siemens Healthineers). TNF (RAF128R, R&D) and IL1b (KAC1211, Invitrogen) were quantified by ELISA. Protein concentrations were standardized to the maximal value per donor.

### Statistical analysis

Statistical analyses were performed using JMP 16 (SAS). Longitudinal comparisons were performed using paired, two-tailed t-tests. Multiple linear regression analyses were performed using log transformed data and adjusted as indicated in the figure legends.

## Results

The clinical characteristics of the study participants have been previously published [11, 20] and are shown in the <u>ESM</u> Table 1. In brief, the eight-week endurance training resulted in significant improvements of the cardiorespiratory fitness quantified as VO_2_peak and of the individual anaerobic threshold as well as in reductions of the BMI and adipose tissue volume (<u>ESM</u> Table 1). As expected from previous studies [21, 22], we observed a large individual variability in the change in peripheral insulin sensitivity (ISI_Mats_), which resulted in an overall non-significant improvement (<u>ESM</u> Table 1). As reported recently, this variance was not due to differences in training volume or intensity [17].

Of the 297 metabolites quantified using our combined CE- and LC-MS-based metabolomics analysis, 85 were significantly changed (median fold change >1.1, p<0.01) after the initial 30 min-cycling bout (Fig. 1 a and <u>ESM</u> Table 2). The majority of metabolites affected by acute exercise showed an increase, with the known exercise-induced metabolites lactate and hypoxanthine and the recently described N-Lactoylphenylalanine (Lac-Phe) exhibiting an increase of more than 300% (Fig. 1a). In contrast, chronic effects of exercise, quantified in the resting state, were more moderate. Only four metabolites were significantly altered by training (median fold change >1.1, p<0.01; Fig. 1 b), all of which were increased. All four were amino acid breakdown products: indole-3-acrylic acid, 3-phenyllactic acid (PLA), 4-hydroxyphenyllactic acid (HPLA), and indolelactic acid (ILA) (Fig. 1 b). Interestingly, the three aromatic lactic acids, PLA, HPLA and ILA, were not only increased after training, but also after acute exercise (labelled in red in Fig. 1 a and b). Among them, PLA showed the most pronounced increases after both acute exercise and training (57% and 20%, respectively, Fig. 1 a and b). All aromatic lactic acids were not only increased after the initial exercise bout but also after the final acute exercise bout performed at the end of the training intervention (Fig. 1 c-e; <0.1 for HPLA). In contrast to the elevated resting concentrations, post-acute exercise levels of PLA, HPLA and ILA were similar before and after the intervention (Fig. 1 c-e).

**Fig. 1:**
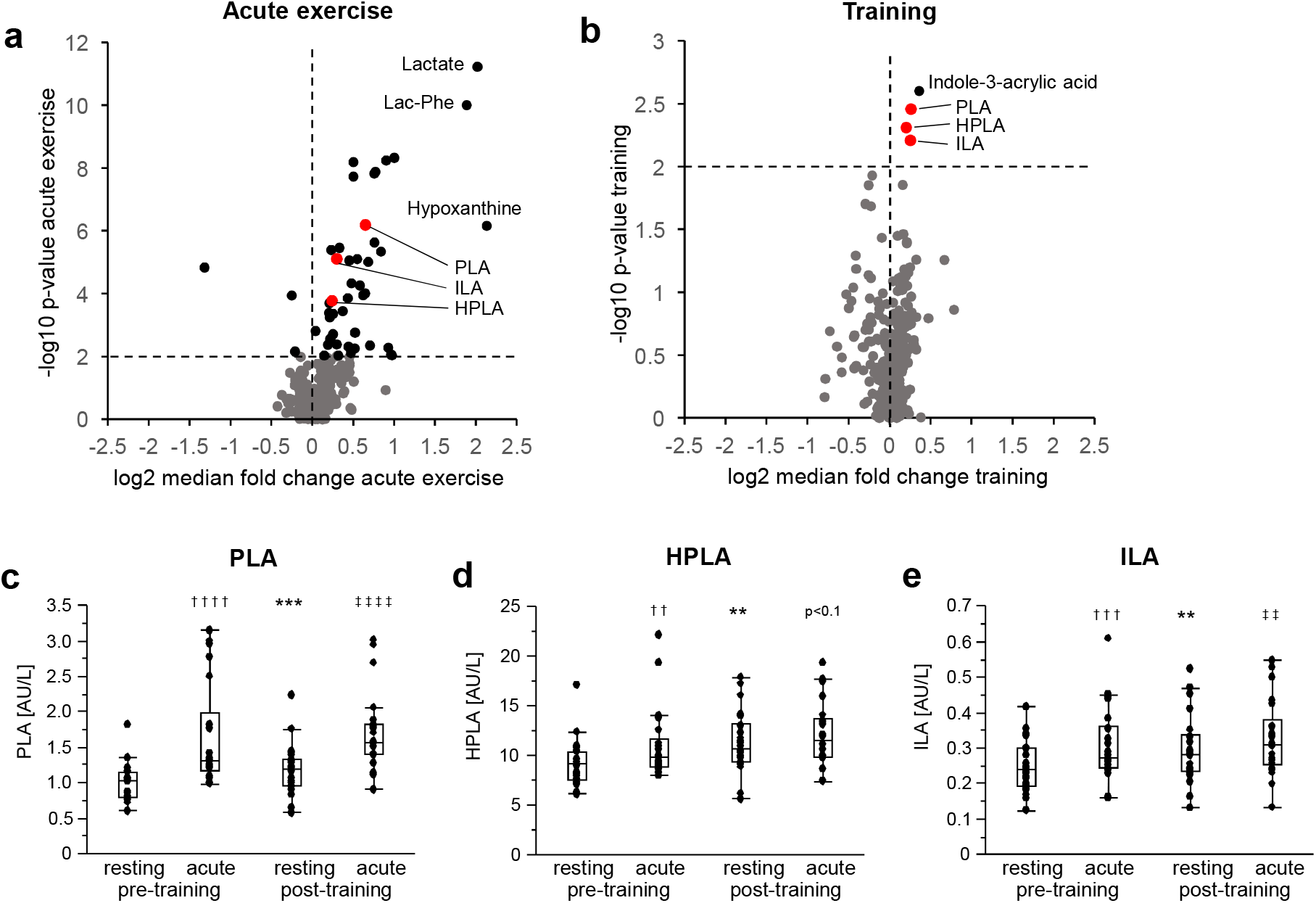
Volcano plots highlighting plasma metabolites changed by (a) acute exercise (pre-training session) and (b) training. Metabolites significantly increased by both acute exercise and training (fold change >1.1, p<0.01) are labelled in red. (c-e): Plasma concentrations of the aromatic lactic acids 3-phenyllactic acid (PLA), 4-hydroxyphenyllactic acid (HPLA) and indolelactic acid (ILA) in the resting state or after a 30-min acute exercise bout performed before (pre-training) or after (post-training) an 8-week supervised endurance exercise intervention (n=22). Timepoints were compared by paired t-tests; \**p*<0.05, \*\**p*<0.01, \*\*\**p*<0.001, ****p<0.0001 for pre- versus post-training, resting; † and ‡ for acute versus resting, pre- and post-training, respectively. A trend (p<0.1) towards an acute exercise-induced increase post-training was identified for HPLA.

In the recent years, aromatic lactic acids received attention as metabolites produced by lactic acid bacteria and present in fermented foods that can be absorbed from the gut in humans [16, 23, 24]. To assess whether the aromatic lactic acids in plasma could also be produced endogenously by skeletal muscle, we investigated samples from a separate study performed in healthy subjects. Here, catheters were placed in femoral vein and artery to quantify metabolite flux over the legs. As expected and previously published [25], we observed a significant efflux of lactate from both legs in the resting, fasting state that was pronouncedly increased for the exercising leg after 60 minutes of one-legged kicking (Fig. 2 a). In a similar manner, PLA and HPLA were released from both legs at baseline (Fig. 2 b, c). There was also a visual increase in the efflux of PLA, HPLA and ILA from the exercising leg at 60 and 120 minutes, however, this effect did not reach statistical significance (Fig. 2 b-d).

**Fig. 2:**
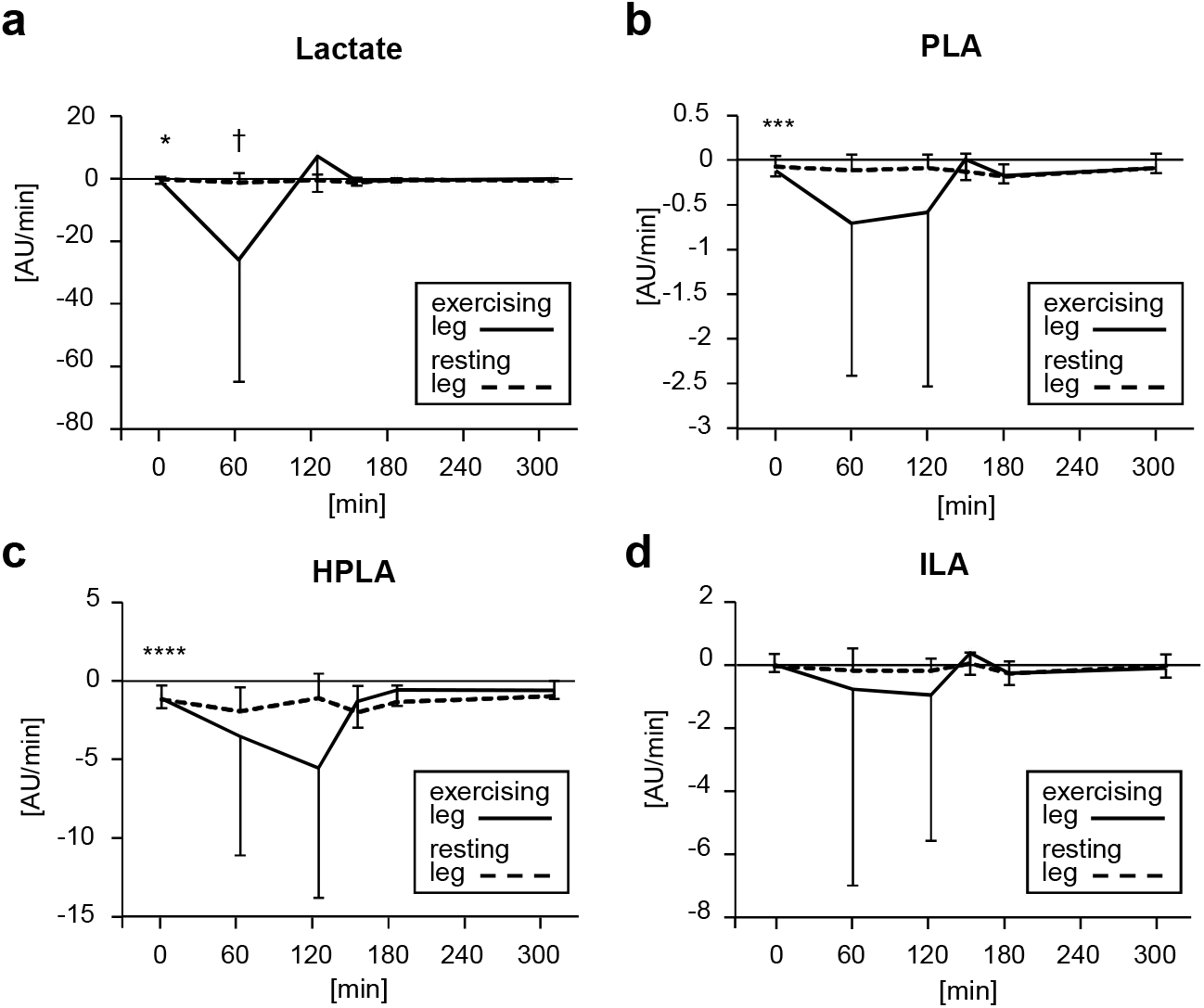
Flux of (a) lactate, (b) 3-phenyllactic acid (PLA), (c) 4-hydroxyphenyllactic acid (HPLA) and (d) indolelactic acid (ILA) from the exercising and resting leg in the exercise study with catheterization of femoral veins and artery. One-legged exercise was performed between minutes 0 and 120. Data are presented as mean ± standard deviation (n=9). \**p*<0.05, ***p<0.001, **** p<0.0001 for flux from both legs according to t-test versus zero. † p<0.05 according to paired t-test resting versus exercising leg. Lactate data have already been published [25].

A recent study demonstrated that oral PLA supplementation counteracts abdominal fat and weight gain in mice fed a high-fat diet [26]. Moreover, plasma levels of PLA have been reported to be lower in subjects with type 2 diabetes compared to healthy controls [27]. We therefore focussed on PLA in our further studies and investigated whether the elevated PLA levels achieved during physical exercise sessions could contribute to individual differences in the reduction of adipose tissue volume or improvement in insulin sensitivity during the eight-week exercise intervention. While the intervention caused a significant reduction in subcutaneous abdominal adipose tissue mass, there was a relevant degree of individual variability (mean change -5%, range -19% to +13%, Supplementary Table S1). Also due to a large degree of heterogeneity, insulin sensitivity was only numerically, but not significantly increased over the whole cohort (mean change +14%, range -44% to +109%, Supplementary Table S1). In support of a potential contribution of exercise-induced PLA to this heterogeneity, its acute increase showed a positive relationship to the change in insulin sensitivity during the intervention (Fig. 3 a, b) and an inverse correlation to the change in subcutaneous adipose tissue over the course of the intervention (Fig. 3 c, b). We did not observe significant correlations of HPLA and ILA with changes in fat mass or insulin sensitivity (data not shown).

**Fig. 3:**
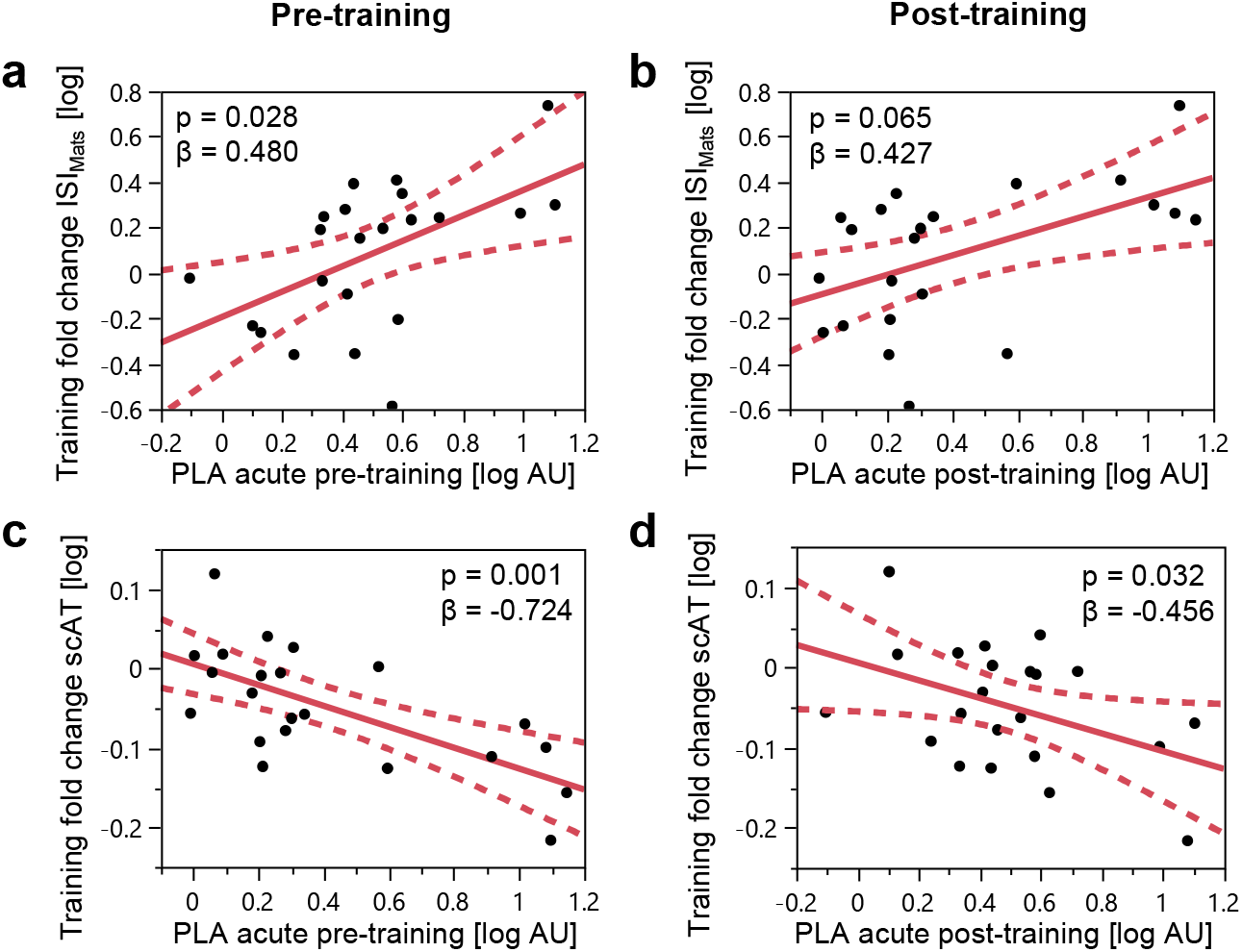
Correlation of the 3-phenyllactic acid (PLA) plasma concentration after acute exercise with the changes in insulin sensitivity (ISI_Mats_) (a, b) and subcutaneous adipose tissue (scAT) mass (c, d) over the 8-week endurance training (n=22). Correlations are shown for the plasma PLA concentrations after both the pre-training (a, c) and post-training acute exercise bout (b, d). P-values and standard β coefficients are shown for the adjusted models (Training fold change scAT: sex, age, pre-training scAT. Training fold change ISI_Mats_: sex, age, BMI, pre-training ISI_Mats_).

Next, we studied the potential function of PLA as a modulator of inflammation [23]. Since human blood has been shown to contain both D- and L-PLA [28], we investigated the effect of both isomers on the inflammatory response. In PBMCs stimulated with LPS, D- and L-PLA counteracted the increase in *IL1B*, *IL6* and *TNF* transcripts and the release of IL1B, IL6 and TNF protein to the same extent and in a concentration- and time-dependent manner (Fig. 4 a-c). Both isomers also inhibited cytokine production induced by the saturated fatty acid palmitate (Fig. 4 d).

**Fig. 4:**
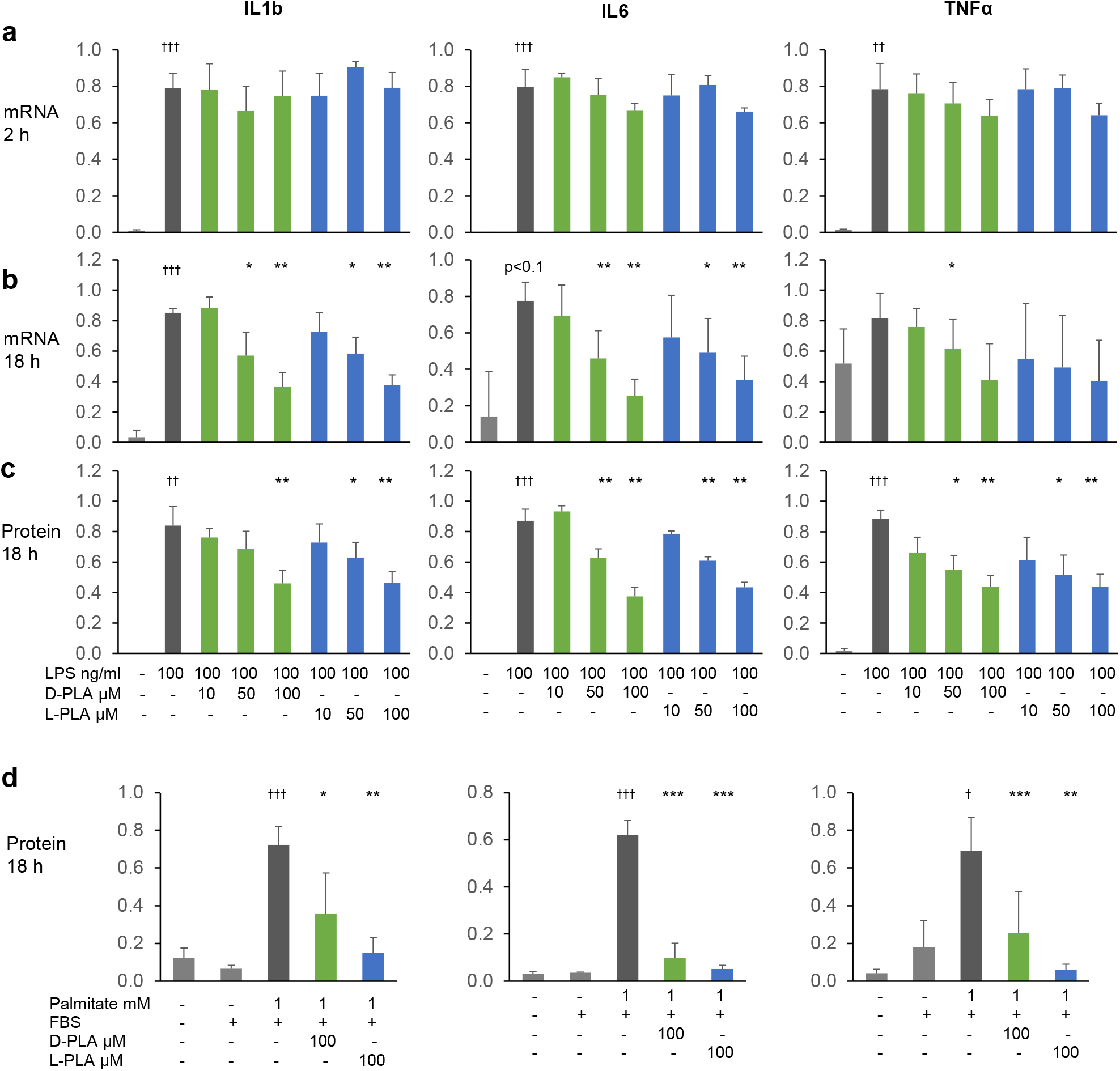
Cytokine mRNA and protein levels in the supernatant of primary blood mononuclear cells (PBMCs) treated with lipopolysaccharide (LPS) or palmitate (in FBS) for 2 h or 18 h after 1 h of preincubation with D- or L-PLA (n=4 donors). mRNA levels were normalized to the reference transcript RPS28 and standardized to the maximal value per donor before calculating means of technical duplicates. Protein concentrations were standardized to the maximum value per donor before calculating means of technical duplicates. Data are presented as mean ± standard deviation and were compared using paired t-tests; † p<0.05, ††† p<0.001 for LPS vs. control and palmitate vs. FBS; \**p*<0.05, \*\**p*<0.01, ***p<0.001 vs. LPS or palmitate.

The hydroxycarboxylic acid receptor (HCAR)3 has recently been described as a receptor for PLA [23]. However, a role of HCAR3 in the effects observed here is unlikely since D-PLA has been reported to activate HCAR3 with a 100-fold higher potency than L-PLA [23] while we observed a similar dose dependency of the two isomers. Moreover, the mRNA levels of *HCAR3* in PBMCs were more than one order of magnitude lower than in neutrophils from the same donors (484±499 vs. 8312±5960 ag/ng total RNA, <u>ESM</u> Fig. 1), which also argues against a major role of HCAR3 in the anti-inflammatory effects observed here.

## Discussion

Exercise-regulated metabolites could play an important and potentially underappreciated role in the beneficial adaptation to physical activity [7–9]. Metabolites with signalling function may be particularly relevant if they are not only acutely elevated after physical exercise, but also at rest in trained subjects. Here, we identified the aromatic lactic acids PLA, HPLA and ILA as a family of metabolites fulfilling these criteria.

Aromatic lactic acids are derivates of the amino acids phenylalanine, tyrosine and tryptophan. Up to now, they mainly received attention as metabolites present in fermented food and produced by gut microbiota that could have beneficial effects on their hosts [23, 24, 26].

However, aromatic lactic acids are also produced by cells of the human body. This becomes apparent in subjects with phenylketonuria where deficiency in the liver enzyme phenylalanine hydroxylase drives major amounts of phenylalanine into transamination to phenylpyruvate, which can further be reduced to PLA [29]. The latter step is catalyzed by lactate dehydrogenase (LDH) isozymes in both humans and bacteria. In humans, LDH is a near-equilibrium enzyme and flux is controlled by increases in its reaction substrates, most importantly pyruvate and NADH, which are increasingly produced by glycolysis during exercise [30]. While the subcellular quantification of NADH and of the NADH/NAD^+^ ratio remain technically challenging [31], *in silico* models indicate that the production of lactate is closely related to the cytosolic NADH/NAD^+^ ratio [32].

Thus, it is at least conceivable that an increase in the cytosolic NADH/NAD^+^ ratio during exercise drives the formation of aromatic lactic acids by LDH along with the production of lactate. In addition, accumulation of protons from increased ATP hydrolysis could promote flux through LDH during exercise [33]. Another factor potentially driving the formation of aromatic lactic acid during exercise may be the availability of free amino acids due to a combination of increased proteolysis and reduced protein synthesis [34].

While we only detected a numerical exercise-induced increase in the efflux of PLA, HPLA, and ILA in our one-leg exercise study with catheterization, the arterial-to venous flux measurements obtained in the resting state are a strong indicator that skeletal muscle contributes to circulating aromatic lactic acids in healthy humans. To our knowledge, no study so far has quantified the contribution of skeletal muscle to the total aromatic lactic acid pool. Our data support the concept that at least under certain conditions, a proportion of circulating PLA can be produced by the human body.

Physical activity is generally health-promoting, but not all individuals benefit equally from exercise-based lifestyle interventions with regard to specific therapeutic goals, such as improved glycaemic control or reduction of adipose tissue mass [2–5]. Prompted by a recent report of a fat-lowering effect of PLA administration in mice [26], we investigated whether the elevated PLA levels achieved during the acute exercise sessions in our study could contribute to individual differences in the response to the training intervention. Indeed, subjects who achieved a more pronounced increase in PLA after acute exercise lost more subcutaneous adipose tissue over the course of the eight weeks of endurance training. To a lesser extent, exercise-induced PLA also correlated with the improvement in insulin sensitivity during the intervention.

We did not observe significant correlations of HPLA and ILA with changes in fat mass or insulin sensitivity and in addition, PLA was the aromatic lactic acid exhibiting the most pronounced increases after both acute exercise and training. Therefore, we focused on PLA in our further mechanistic experiments. While L-PLA is the isomer produced by human cells and also the more abundant form in blood [28], immunomodulatory effects have mainly been attributed to D-PLA produced by bacteria [23]. Thus, we investigated the potential of both isomers as modulators of inflammation. We selected PBMCs as a model system since this leukocyte fraction is enriched in monocytes, a cell type that has previously been shown to be affected by PLA via the receptor HCAR3 [23]. However, D-PLA has a pronouncedly higher affinity for HCAR3 than L-PLA [23] while in the present study, both isomers counteracted the production of IL1B, IL6, and TNF by PBMCs activated with LPS or palmitate to the same degree. Together with our observation that HCAR3 was only expressed at low levels in PBMCs when compared to neutrophils, a likely conclusion is that the anti-inflammatory effects of PLA observed here are HCAR3-independent. Acidification has also been shown to inhibit the activation of monocytes, however, this requires concentrations in the mM range as shown for lactic acid [35, 36]. An alternative mechanism underlying the anti-inflammatory action of PLA could be to decrease ROS production [37]. PLA could also act via peroxisome proliferator-activated receptor (PPAR)-γ, which plays a role both in immune cell and adipocyte function [38] and has been associated with PLA activity in intestinal epithelium and adipocytes [26, 39].

Most certainly, PLA exerts its potential beneficial effects in conjunction with other exercise-induced metabolites. Lactate produced during physical exercise has been suggested to act as a pleiotropic signalling molecule in various cell types and organs and associated with the regulation of energy balance, food intake and thermogenesis [33, 40–42]. However, in experiments where lactate was administered in vivo, at least some of the effects may have been caused by co-administered counterions and osmolality [40]. Similar to Lac-Phe [11, 43], PLA could be a metabolite that mediates anti-obesity and potentially, other beneficial effects that have previously been associated with lactate. With increasing technical advances in metabolomics techniques, additional exercise-induced, lactate-related metabolites with signalling function that have so far been overlooked due to their relatively low abundance may be identified in the future.

One limitation of our study is that it did not include a resting control group. Another limitation is that we were not able to separately quantify the D- and L-isomer, which might have supported the hypothesis that skeletal muscle rather than the microbiome contributed to the bulk of acute exercise-induced increase in aromatic lactic acids. However, it is not possible to unequivocally identify the origin of PLA and other aromatic lactic acids in human blood based on its chirality as gut-colonizing bacterial strains also produce relevant amounts of the L-isomer [28] and humans also express an D-LDH in addition to the predominant L-form isozymes [44]. Importantly, the breakfast served before the acute exercise bout was standardized and contained no fermented products, thus excluding it as a major direct source of aromatic lactic acids. In addition, increased levels of aromatic lactic acids have previously been observed in subjects performing exercise in the fasting state [45, 46].

We did not investigate the mechanism underlying the training-induced increase in aromatic lactic acids. However, increases in faecal concentrations have previously been reported for PLA [47], ILA and HPLA [48] in subjects with obesity participating in endurance exercise interventions. This points towards a contribution of the gut microbiome, which is well-known to be affected by physical exercise [47, 49, 50]. Interestingly, a recent study found that serum ILA and HPLA were increased in both lean and obese subjects following an exercise intervention, whereas faecal levels were increased in the obese cohort only [48]. This suggests that exercise may have diverging effects depending on the obesity status. The same study also reported a correlation of pre-training serum levels of ILA and HPLA with the lean body mass and a lack of correlation between the two metabolites in faeces and serum. This could support a physiologic contribution of muscle tissue to circulating aromatic lactic acids. Further studies are required to quantify what proportion of aromatic lactic acids in the blood originates from the muscle and what from the gut microbiome. This is also relevant since exercise can acutely increase gut permeability [51]. It could be speculated that some of the health benefits of lactic acid bacteria and fermented foods [23, 52] are be related to their ability to produce “exercise-mimicking” metabolites.

Taken together, PLA could be a novel exercise-inducible metabolite that exhibits anti-inflammatory effects and contributes to differences in the individual training response. Since PLA can be produced by commensal bacteria and absorbed from the gut, it is a potential candidate for dietary and microbiome-directed approaches aimed at improving insulin resistance and chronic low-grade inflammation.

## Supporting information

Electronic supplementary material (ESM)

## Acknowledgements

The authors thank all the study participants. The authors are grateful for the excellent technical support provided by Nadine Sanabria Valdés (Institute for Clinical Chemistry and Pathobiochemistry, University Hospital Tübingen). We thank Jakob Schiøler Hansen for his contribution to the leg catheterization study and Prof. Dr. Reinhild Klein for sharing scientific expertise on PBMCs.

## Ethics declarations

### Data availability

Data will only be made available to interested researchers upon reasonable request as far as privacy and consent of research participants are not compromised.

### Authors’ relationships and activities

The authors declare that there are no relationships or activities that might bias, or be perceived to bias, their work.

### Contribution statement

MH designed functional experiments, analyzed and interpreted the data and wrote and edited the manuscript. XZ and CH provided guidance for LC-MS and CE-MS analyses and were responsible for data curation. PP designed and executed the metabolite flux study. ALB, MH, AP and, AN provided scientific guidance and contributed to the discussion. AM designed the intervention study and analyzed anthropometric data. QL performed LC-MS and CE-MS metabolomics analyses. RL provided scientific guidance and experimental design and contributed to the discussion. GX provided scientific guidance and experimental design, contributed to the discussion and reviewed the manuscript. CW designed the intervention study, supervised the whole project, contributed to the discussion, reviewed the manuscript and is the guarantor of the study. All authors approved the final version of the manuscript.

### Prior presentation

A non–peer-reviewed version of this article has been submitted to the medRxiv preprint server on 29 March 2024.

## Abbreviations

PLA: 3-Phenyllactic acid
HPLA: 4-hydroxyphenyllactic acid
CE-QTOF/MS: Capillary electrophoresis time-of-flight mass spectrometry
HCAR3: Hydroxycarboxylic acid receptor 3
ILA: Indolelactic acid
LDH: Lactate dehydrogenase
Lac-Phe: N-lactoyl-phenylalanine
LPS: Lipopolysaccharide
PBMCs: Primary blood mononuclear cells
UHPLC-TOF/MS: Ultra-high-performance liquid chromatography-quadruple-time-of-flight mass spectrometry

## References

1. Karstoft K, Pedersen BK (2016) Exercise and type 2 diabetes: focus on metabolism and inflammation. Immunol Cell Biol 94(2):146–150. 10.1038/icb.2015.101

2. Sparks LM (2017) Exercise training response heterogeneity: physiological and molecular insights. Diabetologia 60(12):2329–2336. 10.1007/s00125-017-4461-6

3. O’Donoghue G, Kennedy A, Andersen GS, et al (2019) Phenotypic Responses to a Lifestyle Intervention Do Not Account for Inter-Individual Variability in Glucose Tolerance for Individuals at High Risk of Type 2 Diabetes. Front Physiol 10:317. 10.3389/fphys.2019.00317

4. Magalhães JP, Hetherington-Rauth M, Júdice PB, et al (2021) Interindividual Variability in Fat Mass Response to a 1-Year Randomized Controlled Trial With Different Exercise Intensities in Type 2 Diabetes: Implications on Glycemic Control and Vascular Function. Front Physiol 12:698971. 10.3389/fphys.2021.698971

5. Solomon TPJ (2018) Sources of Inter-individual Variability in the Therapeutic Response of Blood Glucose Control to Exercise in Type 2 Diabetes: Going Beyond Exercise Dose. Front Physiol 9:896. 10.3389/fphys.2018.00896

6. Pedersen BK, Steensberg A, Fischer C, et al (2003) Searching for the exercise factor: is IL-6 a candidate? J Muscle Res Cell Motil 24(2–3):113–119. 10.1023/a:1026070911202

7. Maurer J, Hoene M, Weigert C (2021) Signals from the Circle: Tricarboxylic Acid Cycle Intermediates as Myometabokines. Metabolites 11(8):474. 10.3390/metabo11080474

8. Yang YR, Kwon K-S (2020) Potential Roles of Exercise-Induced Plasma Metabolites Linking Exercise to Health Benefits. Front Physiol 11:602748. 10.3389/fphys.2020.602748

9. Fernández-Veledo S, Marsal-Beltran A, Vendrell J (2024) Type 2 diabetes and succinate: unmasking an age-old molecule. Diabetologia 67(3):430–442. 10.1007/s00125-023-06063-7

10. Li VL, He Y, Contrepois K, et al (2022) An exercise-inducible metabolite that suppresses feeding and obesity. Nature 606(7915):785–790. 10.1038/s41586-022-04828-5

11. Hoene M, Zhao X, Machann J, et al (2022) Exercise-Induced N-Lactoylphenylalanine Predicts Adipose Tissue Loss during Endurance Training in Overweight and Obese Humans. Metabolites 13(1):15. 10.3390/metabo13010015

12. Tanianskii DA, Jarzebska N, Birkenfeld AL, O’Sullivan JF, Rodionov RN (2019) Beta-Aminoisobutyric Acid as a Novel Regulator of Carbohydrate and Lipid Metabolism. Nutrients 11(3):524. 10.3390/nu11030524

13. Reddy A, Bozi LHM, Yaghi OK, et al (2020) pH-Gated Succinate Secretion Regulates Muscle Remodeling in Response to Exercise. Cell 183(1):62–75.e17. 10.1016/j.cell.2020.08.039

14. Brooks GA, Osmond AD, Arevalo JA, et al (2022) Lactate as a major myokine and exerkine. Nat Rev Endocrinol 18(11):712. 10.1038/s41574-022-00724-0

15. Husted AS, Trauelsen M, Rudenko O, Hjorth SA, Schwartz TW (2017) GPCR-Mediated Signaling of Metabolites. Cell Metab 25(4):777–796. 10.1016/j.cmet.2017.03.008

16. Zhang Y, Chen R, Zhang D, Qi S, Liu Y (2023) Metabolite interactions between host and microbiota during health and disease: Which feeds the other? Biomed Pharmacother 160:114295. 10.1016/j.biopha.2023.114295

17. Hoffmann C, Schneeweiss P, Randrianarisoa E, et al (2020) Response of Mitochondrial Respiration in Adipose Tissue and Muscle to 8 Weeks of Endurance Exercise in Obese Subjects. J Clin Endocrinol Metab 105(11):e4023–e4037. 10.1210/clinem/dgaa571

18. Hansen J, Brandt C, Nielsen AR, et al (2011) Exercise induces a marked increase in plasma follistatin: evidence that follistatin is a contraction-induced hepatokine. Endocrinology 152(1):164–171. 10.1210/en.2010-0868

19. Klein R, Eisenburg J, Weber P, Seibold F, Berg PA (1991) Significance and specificity of antibodies to neutrophils detected by western blotting for the serological diagnosis of primary sclerosing cholangitis. Hepatol Baltim Md 14(6):1147–1152

20. Goj T, Hoene M, Fritsche L, et al (2023) The Acute Cytokine Response to 30-Minute Exercise Bouts Before and After 8-Week Endurance Training in Individuals With Obesity. J Clin Endocrinol Metab 108(4):865–875. 10.1210/clinem/dgac623

21. Böhm A, Weigert C, Staiger H, Häring H-U (2016) Exercise and diabetes: relevance and causes for response variability. Endocrine 51(3):390–401. 10.1007/s12020-015-0792-6

22. Böhm A, Hoffmann C, Irmler M, et al (2016) TGF-β Contributes to Impaired Exercise Response by Suppression of Mitochondrial Key Regulators in Skeletal Muscle. Diabetes 65(10):2849–2861. 10.2337/db15-1723

23. Peters A, Krumbholz P, Jäger E, et al (2019) Metabolites of lactic acid bacteria present in fermented foods are highly potent agonists of human hydroxycarboxylic acid receptor 3. PLoS Genet 15(5):e1008145. 10.1371/journal.pgen.1008145

24. Laursen MF, Sakanaka M, von Burg N, et al (2021) Bifidobacterium species associated with breastfeeding produce aromatic lactic acids in the infant gut. Nat Microbiol 6(11):1367– 1382. 10.1038/s41564-021-00970-4

25. Hu C, Hoene M, Plomgaard P, et al (2019) Muscle-Liver Substrate Fluxes in Exercising Humans and Potential Effects on Hepatic Metabolism. J Clin Endocrinol Metab 105(4):1196– 1209. 10.1210/clinem/dgz266

26. Shelton CD, Sing E, Mo J, et al (2023) An early-life microbiota metabolite protects against obesity by regulating intestinal lipid metabolism. Cell Host Microbe 31(10):1604–1619.e10. 10.1016/j.chom.2023.09.002

27. Zhao Q, Zhang A, Zong W, et al (2017) Exploring potential biomarkers and determining the metabolic mechanism of type 2 diabetes mellitus using liquid chromatography coupled to high-resolution mass spectrometry. RSC Adv 7(70):44186–44198. 10.1039/C7RA05722A

28. Oezguen N, Yılmaz V, Horvath TD, et al (2022) Serum 3-phenyllactic acid level is reduced in benign multiple sclerosis and is associated with effector B cell ratios. Mult Scler Relat Disord 68:104239. 10.1016/j.msard.2022.104239

29. Alsharhan H, Ficicioglu C (2020) Disorders of phenylalanine and tyrosine metabolism. Transl Sci Rare Dis 5(1–2):3–58. 10.3233/TRD-200049

30. Spriet LL, Howlett RA, Heigenhauser GJ (2000) An enzymatic approach to lactate production in human skeletal muscle during exercise. Med Sci Sports Exerc 32(4):756–763. 10.1097/00005768-200004000-00007

31. White AT, Schenk S (2012) NAD+/NADH and skeletal muscle mitochondrial adaptations to exercise. Am J Physiol-Endocrinol Metab 303(3):E308–E321. 10.1152/ajpendo.00054.2012

32. Li Y, Dash RK, Kim J, Saidel GM, Cabrera ME (2009) Role of NADH/NAD+ transport activity and glycogen store on skeletal muscle energy metabolism during exercise: in silico studies. Am J Physiol-Cell Physiol 296(1):C25–C46. 10.1152/ajpcell.00094.2008

33. Nalbandian M, Takeda M (2016) Lactate as a Signaling Molecule That Regulates Exercise-Induced Adaptations. Biology 5(4):38. 10.3390/biology5040038

34. Poortmans JR, Carpentier A, Pereira-Lancha LO, Lancha A (2012) Protein turnover, amino acid requirements and recommendations for athletes and active populations. Braz J Med Biol Res 45(10):875–890. 10.1590/S0100-879X2012007500096

35. Caslin HL, Abebayehu D, Pinette JA, Ryan JJ (2021) Lactate Is a Metabolic Mediator That Shapes Immune Cell Fate and Function. Front Physiol 12:688485. 10.3389/fphys.2021.688485

36. Schenz J, Heilig L, Lohse T, et al (2021) Extracellular Lactate Acts as a Metabolic Checkpoint and Shapes Monocyte Function Time Dependently. Front Immunol 12:729209. 10.3389/fimmu.2021.729209

37. Beloborodova N, Bairamov I, Olenin A, Shubina V, Teplova V, Fedotcheva N (2012) Effect of phenolic acids of microbial origin on production of reactive oxygen species in mitochondria and neutrophils. J Biomed Sci 19(1):89. 10.1186/1423-0127-19-89

38. Hernandez-Quiles M, Broekema MF, Kalkhoven E (2021) PPARgamma in Metabolism, Immunity, and Cancer: Unified and Diverse Mechanisms of Action. Front Endocrinol 12:624112. 10.3389/fendo.2021.624112

39. Ilavenil S, Kim DH, Arasu MV, Srigopalram S, Sivanesan R, Choi KC (2015) Phenyllactic Acid from Lactobacillus plantarum Promotes Adipogenic Activity in 3T3-L1 Adipocyte via Up-Regulation of PPAR-γ2. Molecules 20(8):15359. 10.3390/molecules200815359

40. Lund J, Breum AW, Gil C, et al (2023) The anorectic and thermogenic effects of pharmacological lactate in male mice are confounded by treatment osmolarity and co-administered counterions. Nat Metab 5(4):677–698. 10.1038/s42255-023-00780-4

41. Carrière A, Lagarde D, Jeanson Y, et al (2020) The emerging roles of lactate as a redox substrate and signaling molecule in adipose tissues. J Physiol Biochem 76(2):241–250. 10.1007/s13105-019-00723-2

42. Lezi E, Lu J, Selfridge JE, Burns JM, Swerdlow RH (2013) Lactate Administration Reproduces Specific Brain and Liver Exercise-Related Changes. J Neurochem 127(1):91–100. 10.1111/jnc.12394

43. Lund J, Clemmensen C, Schwartz TW (2022) Outrunning obesity with Lac-Phe? Cell Metab 34(8):1085–1087. 10.1016/j.cmet.2022.07.007

44. Flick MJ, Konieczny SF (2002) Identification of putative mammalian D-lactate dehydrogenase enzymes. Biochem Biophys Res Commun 295(4):910–916. 10.1016/s0006-291x(02)00768-4

45. Morville T, Sahl RE, Moritz T, Helge JW, Clemmensen C (2020) Plasma Metabolome Profiling of Resistance Exercise and Endurance Exercise in Humans. Cell Rep 33(13):108554. 10.1016/j.celrep.2020.108554

46. Grapov D, Fiehn O, Campbell C, et al (2019) Exercise plasma metabolomics and xenometabolomics in obese, sedentary, insulin-resistant women: impact of a fitness and weight loss intervention. Am J Physiol Endocrinol Metab 317(6):E999–E1014. 10.1152/ajpendo.00091.2019

47. Hintikka JE, Ahtiainen JP, Permi P, Jalkanen S, Lehtonen M, Pekkala S (2023) Aerobic exercise training and gut microbiome-associated metabolic shifts in women with overweight: a multi-omic study. Sci Rep 13(1):1–12. 10.1038/s41598-023-38357-6

48. Kasperek MC, Mailing L, Piccolo BD, et al (2023) Exercise training modifies xenometabolites in gut and circulation of lean and obese adults. Physiol Rep 11(6):e15638. 10.14814/phy2.15638

49. Monda V, Villano I, Messina A, et al (2017) Exercise Modifies the Gut Microbiota with Positive Health Effects. Oxid Med Cell Longev 2017:3831972. 10.1155/2017/3831972

50. Boytar AN, Skinner TL, Wallen RE, Jenkins DG, Dekker Nitert M (2023) The Effect of Exercise Prescription on the Human Gut Microbiota and Comparison between Clinical and Apparently Healthy Populations: A Systematic Review. Nutrients 15(6):1534. 10.3390/nu15061534

51. Keirns BH, Koemel NA, Sciarrillo CM, Anderson KL, Emerson SR (2020) Exercise and intestinal permeability: another form of exercise-induced hormesis? Am J Physiol Gastrointest Liver Physiol 319(4):G512–G518. 10.1152/ajpgi.00232.2020

52. Lee M, Kim D, Chang JY (2023) Metabolites of Kimchi Lactic Acid Bacteria, Indole-3-Lactic Acid, Phenyllactic Acid, and Leucic Acid, Inhibit Obesity-Related Inflammation in Human Mesenchymal Stem Cells. J Microbiol Biotechnol 34(3):1–8. 10.4014/jmb.2308.08015

