## Supplementary material for "Phenyllactic Acid is Physiologically Released from Skeletal Muscle and Contributes to the Beneficial Effects of Physical Exercise in Humans": Electronic supplementary material (ESM)

**Table of contents:**

ESM Methods

ESM Tables 1-2

ESM Figure 1

#### ESM Methods

**CE-MS-based profiling analysis of plasma metabolites**

Sample preparation for capillary electrophoresis time-of-flight mass spectrometry (CE-TOF/MS) was performed as described in our previous study [1]. Briefly, 50 µl EDTA plasma were mixed with 450 µL MeOH containing internal standards (10 µM of L-methionine sulfone and D-camphor-10-sulfonic acid sodium salt, 0.9 µg/mL of L-alanine-3,3,3-d3, 0.9 µg/mL of succinic acid-13C4, and 0.7 µg/mL of cholic acid-2,2,4,4-d4), followed by the addition of 500 µL of $CHCl_3$ and 200 µL of water. After vortexing and centrifugation to form a 2-phase system, the upper layer was centrifugally filtered through a 5-kDa cutoff filter (HMT, Japan) to remove residual proteins. The filtrate was lyophilized and stored for later analysis. Prior to CE-TOF/MS metabolomics analysis, lyophilized samples were preconditioned in Milli-Q water containing the internal standards 1 mM 3-aminopyrrolidine dihydrochloride (Aldrich, USA), N,N-Diethyl-2-phenylacetamide (Wako, Japan), trimesic acid (Wako, Japan), and disodium 3-hydroxynaphthalene-2,7-disulfonate (Wako, Japan) for migration time correction.

CE-TOF/MS analysis was performed on a CE (G7100A, Agilent)-TOF/MS (G6224A, Agilent) system equipped with an ESI-MS sprayer kit (G1607A, Agilent), an 1260 ISO pump (G1310B, Agilent) and a minichiller (Huber, Germany) as described previously [1]. A fused-silica capillary (80 cm × 50 µm i.d.) was used for CE separation. The capillary temperature was maintained at 20 °C. The sample tray temperature was set at 5°C controlled by the minichiller. The CE-TOF/MS coupling was realized by a coaxial sheath liquid interface. The sheath liquid containing MeOH/water (1:1, v/v) and 0.1 µM hexakis (2, 2-difluoroethoxy) phosphazene was delivered at 10 µL/min. CE-TOF/MS data acquisition of plasma samples was carried out in both cation and anion modes [2].

**LC-MS-based profiling analysis of plasma metabolites**

Sample preparation for liquid chromatography-mass spectrometry (LC-MS) was performed by deproteinizing 150 µl EDTA with 750 µl MeOH containing the internal standards L-methionine sulfone, D-camphor-10-sulfonic acid sodium salt, leucine-d3, phenylalanine-d5, tryptophane-d5, succinic acid-13C4, cholic acid-d4 (all from Sigma-Aldrich, Steinheim, Germany), and free fatty acid 12:0-d4 and carnitines C0-d3 C2:0-d3 C10:0-d3 and C16:0-d3 (all ten Brink laboratories, Amsterdam, Netherlands). After vortexing and centrifugation (16,000 xg, 20 min, 4 °C), supernatants were lyophilized and stored for later analysis.

Ultra-high-performance liquid chromatography-quadruple-time-of-flight mass spectrometry (UHPLC-QTOF/MS) analysis was performed on a Thermo Scientific Vanquish UHPLC coupled to a Q Exactive mass spectrometer (Thermo). For position ion scan mode, an ACQUITY UPLC $C_8$ column (50 mm × 2.1 mm, 1.7 µm) (Waters, Milford, MA) was used for LC separation. Pure water containing 0.1% formic acid (A) and acetonitrile containing 0.1% formic acid (B) were used as binary mobile phase composition. Gradient elution started with 5% B and kept for 0.5 min. After having been increased to 40% in 1.5 min, B was further increased to 100% in 6 min and kept isocratic for 2 min. Finally, B was decreased back to 5% in 0.1 min and maintained for additional 1.9 min. For negative ion scan mode, an ACQUITY UPLC HSS T3 column (50 mm × 2.1 mm, 1.8 µm) (Waters, Milford, MA) was used for LC separation. The binary mobile phase consisted of pure water (A) and 95% (v/v) methanol/water (B), both containing 6.5 mM ammonium bicarbonate. The initial gradient consisting of 2% B was held for 0.5 min and then increased to 40% in 2 min, to 100% in 6 min and kept for 2 min. Finally, B was rapidly decreased to 5%

within 0.1 min and kept for 1.9 min for column re-equilibration. Flow rate and column temperature in both ionization modes were set at 0.4 mL/min and 60 °C, respectively.

MS condition was set as follows: sheath gas flow rate, 45 arb; Aux gas flow rate, 10 arb; spray voltage, 3.5 kV and - 3.0 kV for positive and negative ion scan modes, respectively; capillary temp: 320 °C; Aux gas heater temp, 350 °C; resolution 14 0,000. Full MS ion scan mode with scan range of 70 - 1050 m/z was used for both positive and negative ion scan.

**References**

1. Hu C, Hoene M, Plomgaard P, et al (2019) Muscle-Liver Substrate Fluxes in Exercising Humans and Potential Effects on Hepatic Metabolism. J Clin Endocrinol Metab 105(4):1196–1209. https://doi.org/10.1210/clinem/dgz266

2. Zeng J, Kuang H, Hu C, et al (2013) Effect of bisphenol A on rat metabolic profiling studied by using capillary electrophoresis time-of-flight mass spectrometry. Environ Sci Technol 47(13):7457–7465. https://doi.org/10.1021/es400490f

### ESM Tables

**Table 1:** Anthropometric, fitness and metabolic data

| | **Pre-training** | **Post-training** | **Fold change** | ***p*-Value** |
|---|---|---|---|---|
| Sex | 14 female, 8 male | | | - |
| Age [years] | 30 ± 9 (19–59) | | | - |
| BMI [kg/m$^2$] | 31.7 ± 4.5 (27.5–45.5) | 31.3 ± 4.7 (26.3–45.2) | 0.99 ± 0.02 (0.95–1.04) | < 0.01 |
| $VO_2peak_{ergo}$ [mL/(kg x min)] | 25 ± 4 (18–32) | 27 ± 5 (16–35) | 1.07 ± 0.14 (0.75–1.35) | < 0.05 |
| $IAT_{ergo}$, W/kg | 1.1 ± 0.2 (0.8–1.6) | 1.3 ± 0.3 (0.9–1.9) | 1.19 ± 0.12 (0.95–1.40) | < 0.001 |
| $ISI_{Mats}$ | 8.4 ± 5.0 (3.1–27.0) | 8.9 ± 4.3 (4.5–21.4) | 1.14 ± 0.35 (0.56–2.09) | > 0.1 |
| Glucose fasting [mmol/L] | 5.09 ± 0.40 (4.61–6.00) | 5.02 ± 0.40 (4.33–5.61) | 0.99 ± 0.06 (0.87–1.08) | > 0.1 |
| Subcutaneous abdominal adipose tissue [L] | 15.3 ± 5.9 (8.4–32.2) | 14.7 ± 6.1 (7.2–33.1) | 0.95 ± 0.07 (0.81–1.13) | < 0.01 |
| Visceral adipose tissue [L] | 3.53 ± 1.65 (0.81–7.26) | 3.38 ± 1.57 (0.94–6.68) | 0.97 ± 0.10 (0.72–1.16) | < 0.05 |

*IAT, individual anaerobic threshold. N = 22, $VO_2peak$ N = 21; mean ± SD (range of values); paired Student's t-test.*

**Table 2:** List of identfied metabolites

| Metabolite | Detection method | Identification method | Acute exercise | | Training | |
|---|---|---|---|---|---|---|
| | | | Median fold change | p-value | Median fold change | p-value |
| 1,7-Dimethylxanthine | LC | standard | 0.93 | 0.0332 | 0.67 | 0.4338 |
| 2-Hydroxy-3-methylbutyric acid | LC | LC-MS database tr,MS1,MS2 | 1.17 | 0.0000 | 1.07 | 0.1826 |
| 3-Indolepropionic acid | LC | standard | 1.11 | 0.0094 | 1.08 | 0.1615 |
| 3-Phenyllactic acid (PLA) | LC | standard | 1.57 | 0.0000 | 1.20 | 0.0035 |
| Cholic acid | LC | standard | 0.40 | 0.0000 | 0.74 | 0.0915 |
| Caffeine | LC | standard | 0.86 | 0.0071 | 0.60 | 0.2035 |
| Carnitine | LC | standard | 1.03 | 0.0016 | 0.94 | 0.1021 |
| Carnitine C10_0 | LC | standard | 0.90 | 0.9700 | 1.22 | 0.2537 |
| Carnitine C10_1 | LC | LC-MS database tr,MS1,MS2 | 1.18 | 0.0385 | 1.14 | 0.1889 |
| Carnitine C10_2 | LC | LC-MS database tr,MS1,MS2 | 1.20 | 0.0475 | 1.23 | 0.2625 |
| Carnitine C12_0 | LC | LC-MS database tr,MS1,MS2 | 0.94 | 0.8045 | 1.15 | 0.6525 |
| Carnitine C12_1 | LC | LC-MS database tr,MS1,MS2 | 0.89 | 0.7237 | 1.03 | 0.6259 |
| Carnitine C14_0 | LC | LC-MS database tr,MS1,MS2 | 1.06 | 0.4991 | 1.11 | 0.4128 |
| Carnitine C14_1 | LC | LC-MS database tr,MS1,MS2 | 0.91 | 0.3739 | 1.13 | 0.6152 |
| Carnitine C14_2 | LC | LC-MS database tr,MS1,MS2 | 1.00 | 0.8591 | 1.09 | 0.2078 |
| Carnitine C14_3 | LC | LC-MS database tr,MS1,MS2 | 0.80 | 0.6257 | 1.09 | 0.4056 |
| Carnitine C16_0 | LC | standard | 1.07 | 0.0477 | 1.01 | 0.8300 |
| Carnitine C16_1-1 | LC | LC-MS database tr,MS1,MS2 | 1.06 | 0.8750 | 1.06 | 0.4577 |
| Carnitine C16_1-2 | LC | LC-MS database tr,MS1,MS2 | 1.05 | 0.3481 | 1.10 | 0.2904 |
| Carnitine C16_2 | LC | LC-MS database tr,MS1,MS2 | 0.91 | 0.5707 | 1.06 | 0.5406 |
| Carnitine C18_0 | LC | LC-MS database tr,MS1,MS2 | 1.11 | 0.1619 | 0.94 | 0.4593 |
| Carnitine C18_1 | LC | LC-MS database tr,MS1,MS2 | 1.27 | 0.0500 | 1.20 | 0.1081 |
| Carnitine C18_2 | LC | LC-MS database tr,MS1,MS2 | 1.42 | 0.0642 | 1.25 | 0.2860 |
| Carnitine C18_3 | LC | LC-MS database tr,MS1,MS2 | 1.28 | 0.0192 | 1.23 | 0.2404 |
| Carnitine C2_0 | LC | standard | 1.46 | 0.0000 | 1.06 | 0.4164 |
| Carnitine C3_0 | LC | LC-MS database tr,MS1,MS2 | 1.86 | 0.1191 | 0.95 | 0.1646 |
| Carnitine C4_0 | LC | standard | 1.39 | 0.0077 | 0.95 | 0.3947 |
| Carnitine C4_0-2 | LC | LC-MS database tr,MS1,MS2 | 1.37 | 0.0326 | 0.81 | 0.0198 |
| Carnitine C5_0 | LC | LC-MS database tr,MS1,MS2 | 1.08 | 0.0274 | 0.84 | 0.0732 |
| Carnitine C5_0-2 | LC | LC-MS database tr,MS1,MS2 | 1.07 | 0.2641 | 0.94 | 0.6005 |
| Carnitine C5_1 | LC | LC-MS database tr,MS1,MS2 | 1.15 | 0.1097 | 0.83 | 0.7431 |

| | | | | | | |
|---|---|---|---|---|---|---|
| Carnitine C6_0 | LC | standard | 1.14 | 0.0663 | 1.06 | 0.5999 |
| Carnitine C8_0 | LC | LC-MS database tr,MS1,MS2 | 0.93 | 0.9511 | 1.12 | 0.3938 |
| Carnitine C8_1-1 | LC | LC-MS database tr,MS1,MS2 | 1.13 | 0.0251 | 1.17 | 0.1285 |
| Carnitine C8_1-2 | LC | LC-MS database tr,MS1,MS2 | 1.14 | 0.0043 | 1.01 | 0.6934 |
| Choline | LC | LC-MS database tr,MS1,MS2 | 0.98 | 0.6354 | 1.19 | 0.0635 |
| creatinine | LC | standard | 1.06 | 0.2783 | 0.96 | 0.2182 |
| DCA | LC | standard | 0.82 | 0.2596 | 0.74 | 0.4027 |
| delta-Valerolactam | LC | LC-MS database tr,MS1,MS2 | 1.23 | 0.3302 | 1.00 | 0.5766 |
| Dl-P-Hydroxyphenyl lactic acid (HPLA) | LC | LC-MS database tr,MS1,MS2 | 1.18 | 0.0002 | 1.15 | 0.0049 |
| fatty amide C16_0 | LC | standard | 1.30 | 0.0245 | 1.18 | 0.9284 |
| fatty amide C18_0 | LC | standard | 1.37 | 0.0166 | 1.13 | 0.6436 |
| fatty amide C18_1 | LC | LC-MS database tr,MS1,MS2 | 1.08 | 0.1564 | 1.11 | 0.9157 |
| fatty amide C20_1 | LC | LC-MS database tr,MS1,MS2 | 1.14 | 0.0535 | 1.04 | 0.3101 |
| fatty amide C22_0 | LC | LC-MS database tr,MS1,MS2 | 1.10 | 0.1543 | 0.99 | 0.5031 |
| fatty amide C22_1 | LC | LC-MS database tr,MS1,MS2 | 1.05 | 0.1497 | 0.95 | 0.3777 |
| fatty amide C22_2 | LC | LC-MS database tr,MS1,MS2 | 1.11 | 0.0851 | 0.98 | 0.6562 |
| FFA 10_0 | LC | LC-MS database tr,MS1,MS2 | 0.84 | 0.5737 | 1.07 | 0.7870 |
| FFA 11_0 | LC | LC-MS database tr,MS1,MS2 | 1.07 | 0.7766 | 1.08 | 0.3241 |
| FFA 12_0 | LC | LC-MS database tr,MS1,MS2 | 1.15 | 0.0279 | 1.04 | 0.6446 |
| FFA 13_0 | LC | LC-MS database tr,MS1,MS2 | 1.17 | 0.1515 | 0.99 | 0.5100 |
| FFA 14_0 | LC | standard | 0.96 | 0.7735 | 1.10 | 0.3680 |
| FFA 14_1 | LC | LC-MS database tr,MS1,MS2 | 0.98 | 0.5528 | 1.16 | 0.1813 |
| FFA 15_0 | LC | LC-MS database tr,MS1,MS2 | 0.92 | 0.9055 | 1.15 | 0.1800 |
| FFA 16_0 | LC | standard | 0.94 | 0.4948 | 1.17 | 0.3623 |
| FFA 16_1 | LC | standard | 0.84 | 0.2329 | 1.10 | 0.1525 |
| FFA 16_2 | LC | LC-MS database tr,MS1,MS2 | 1.09 | 0.4248 | 1.10 | 0.0758 |
| FFA 16_3 | LC | LC-MS database tr,MS1,MS2 | 1.05 | 0.2065 | 1.13 | 0.0751 |
| FFA 17_0 | LC | LC-MS database tr,MS1,MS2 | 0.96 | 0.3404 | 1.03 | 0.4726 |
| FFA 17_1 | LC | LC-MS database tr,MS1,MS2 | 0.82 | 0.2703 | 1.16 | 0.2311 |
| FFA 17_2 | LC | LC-MS database tr,MS1,MS2 | 0.96 | 0.6952 | 1.13 | 0.0657 |
| FFA 18_0 | LC | standard | 0.95 | 0.3627 | 1.01 | 0.7880 |
| FFA 18_1 | LC | standard | 0.77 | 0.1710 | 1.09 | 0.2242 |
| FFA 18_2 | LC | standard | 0.83 | 0.2972 | 1.09 | 0.0910 |
| FFA 18_3 | LC | LC-MS database tr,MS1,MS2 | 1.00 | 0.8158 | 1.08 | 0.2632 |

| | | | | | | |
|---|---|---|---|---|---|---|
| FFA 18_4 | LC | LC-MS database tr,MS1,MS2 | 1.02 | 0.8279 | 1.00 | 0.3854 |
| FFA 19_0 | LC | LC-MS database tr,MS1,MS2 | 0.96 | 0.3032 | 0.96 | 0.3654 |
| FFA 19_1 | LC | LC-MS database tr,MS1,MS2 | 0.85 | 0.2584 | 1.08 | 0.4140 |
| FFA 20_0 | LC | LC-MS database tr,MS1,MS2 | 0.96 | 0.7023 | 0.95 | 0.3532 |
| FFA 20_1 | LC | LC-MS database tr,MS1,MS2 | 0.80 | 0.2332 | 1.20 | 0.2747 |
| FFA 20_2 | LC | LC-MS database tr,MS1,MS2 | 0.88 | 0.2386 | 1.11 | 0.1744 |
| FFA 20_3 | LC | LC-MS database tr,MS1,MS2 | 0.82 | 0.2239 | 1.13 | 0.2697 |
| FFA 20_4 | LC | standard | 0.82 | 0.3116 | 1.13 | 0.1543 |
| FFA 20_5 | LC | LC-MS database tr,MS1,MS2 | 0.90 | 0.5707 | 1.05 | 0.7271 |
| FFA 22_0 | LC | LC-MS database tr,MS1,MS2 | 0.98 | 0.7495 | 1.03 | 0.7842 |
| FFA 22_1 | LC | LC-MS database tr,MS1,MS2 | 1.03 | 0.8052 | 1.09 | 0.9705 |
| FFA 22_2 | LC | LC-MS database tr,MS1,MS2 | 1.12 | 0.9651 | 1.11 | 0.4301 |
| FFA 22_3 | LC | LC-MS database tr,MS1,MS2 | 1.09 | 0.7351 | 1.25 | 0.1574 |
| FFA 22_4 | LC | LC-MS database tr,MS1,MS2 | 0.82 | 0.2660 | 1.11 | 0.2028 |
| FFA 22_5 | LC | LC-MS database tr,MS1,MS2 | 0.90 | 0.4219 | 1.21 | 0.1441 |
| FFA 22_6 | LC | LC-MS database tr,MS1,MS2 | 0.90 | 0.2517 | 1.25 | 0.0550 |
| FFA 23_0 | LC | LC-MS database tr,MS1,MS2 | 1.02 | 0.3819 | 1.03 | 0.5371 |
| FFA 24_0 | LC | LC-MS database tr,MS1,MS2 | 0.97 | 0.9869 | 1.06 | 0.9989 |
| FFA 24_1 | LC | LC-MS database tr,MS1,MS2 | 1.01 | 0.6176 | 1.04 | 0.7404 |
| FFA 9_0 | LC | LC-MS database tr,MS1,MS2 | 1.03 | 0.4033 | 1.06 | 0.3764 |
| GCA | LC | standard | 0.83 | 0.5626 | 0.64 | 0.2704 |
| GCDCA | LC | standard | 1.31 | 0.1309 | 0.81 | 0.1992 |
| GCDCA-glucuronide | LC | standard | 0.91 | 0.7016 | 0.75 | 0.0511 |
| GCDCS | LC | standard | 0.94 | 0.3772 | 1.18 | 0.2855 |
| GDCA | LC | standard | 1.08 | 0.5558 | 0.90 | 0.2343 |
| GDCS | LC | standard | 0.92 | 0.8660 | 1.04 | 0.1488 |
| GLCA | LC | standard | 1.91 | 0.0053 | 1.00 | 0.2629 |
| GLCAS | LC | standard | 1.06 | 0.0505 | 0.97 | 0.6481 |
| GUDCA | LC | standard | 1.00 | 0.7608 | 1.05 | 0.7517 |
| GUDCS | LC | standard | 1.00 | 0.8952 | 1.04 | 0.9443 |
| Hippuric acid | LC | standard | 1.22 | 0.0282 | 0.74 | 0.2196 |
| Indole-3-acrylic acid | LC | LC-MS database tr,MS1,MS2 | 0.99 | 0.9876 | 1.28 | 0.0025 |
| Indolelactic acid (ILA) | LC | standard | 1.23 | 0.0000 | 1.19 | 0.0062 |
| N-Lactoylphenylalanine (Lac-Phe) | LC | standard | 3.70 | 0.0000 | 0.98 | 0.7501 |

| | | | | | | |
|---|---|---|---|---|---|---|
| LPC 14_0 sn-1 | LC | standard | 1.03 | 0.6614 | 0.97 | 0.2649 |
| LPC 14_0 sn-2 | LC | LC-MS database tr,MS1,MS2 | 0.92 | 0.7969 | 0.84 | 0.1982 |
| LPC 15_0 sn-1 | LC | LC-MS database tr,MS1,MS2 | 1.03 | 0.5313 | 1.03 | 0.4641 |
| LPC 15_0 sn-2 | LC | LC-MS database tr,MS1,MS2 | 1.04 | 0.9253 | 0.99 | 0.8653 |
| LPC 16_0 sn-1 | LC | standard | 0.98 | 0.4075 | 1.01 | 0.9568 |
| LPC 16_0 sn-2 | LC | LC-MS database tr,MS1,MS2 | 0.94 | 0.0974 | 1.00 | 0.7144 |
| LPC 16_1 sn-1 | LC | LC-MS database tr,MS1,MS2 | 1.01 | 0.4079 | 0.85 | 0.1585 |
| LPC 16_1 sn-2 | LC | LC-MS database tr,MS1,MS2 | 1.09 | 0.9998 | 0.74 | 0.2264 |
| LPC 17_0 | LC | LC-MS database tr,MS1,MS2 | 0.95 | 0.1885 | 1.01 | 0.3510 |
| LPC 18_0 sn-1 | LC | standard | 0.91 | 0.1629 | 1.04 | 0.4437 |
| LPC 18_0 sn-2 | LC | LC-MS database tr,MS1,MS28 | 0.96 | 0.1280 | 1.09 | 0.5330 |
| LPC 18_1 sn-1 | LC | standard | 0.96 | 0.0798 | 0.96 | 0.7219 |
| LPC 18_1 sn-2 | LC | LC-MS database tr,MS1,MS2 | 0.91 | 0.0289 | 0.94 | 0.7173 |
| LPC 18_2 sn-1 | LC | LC-MS database tr,MS1,MS2 | 0.91 | 0.1256 | 1.03 | 0.7832 |
| LPC 18_2 sn-2 | LC | LC-MS database tr,MS1,MS2 | 0.89 | 0.0415 | 0.96 | 0.6153 |
| LPC 18_3 | LC | LC-MS database tr,MS1,MS2 | 0.83 | 0.0331 | 0.71 | 0.1335 |
| LPC 20_0 | LC | LC-MS database tr,MS1,MS2 | 1.03 | 0.9731 | 1.07 | 0.1621 |
| LPC 20_1 | LC | LC-MS database tr,MS1,MS2 | 0.90 | 0.0695 | 1.00 | 0.1715 |
| LPC 20_2 | LC | LC-MS database tr,MS1,MS2 | 0.91 | 0.0294 | 0.95 | 0.6329 |
| LPC 20_3 | LC | LC-MS database tr,MS1,MS2 | 0.90 | 0.0518 | 0.95 | 0.6700 |
| LPC 20_4 | LC | LC-MS database tr,MS1,MS2 | 0.91 | 0.0883 | 0.89 | 0.9580 |
| LPC 20_5 | LC | LC-MS database tr,MS1,MS2 | 0.90 | 0.1608 | 0.80 | 0.2727 |
| LPC 22_0 | LC | LC-MS database tr,MS1,MS2 | 0.94 | 0.9032 | 1.01 | 0.9239 |
| LPC 22_1 | LC | LC-MS database tr,MS1,MS2 | 0.92 | 0.0795 | 1.07 | 0.1622 |
| LPC 22_4 | LC | LC-MS database tr,MS1,MS2 | 0.88 | 0.0556 | 0.91 | 0.6632 |
| LPC 22_5 | LC | LC-MS database tr,MS1,MS2 | 0.87 | 0.0512 | 0.90 | 0.3976 |
| LPC 22_6 | LC | LC-MS database tr,MS1,MS2 | 0.90 | 0.1030 | 0.92 | 0.9983 |
| LPC 24_0 | LC | LC-MS database tr,MS1,MS2 | 1.11 | 0.0228 | 1.07 | 0.0979 |
| LPC O-16_0 | LC | LC-MS database tr,MS1,MS2 | 0.95 | 0.3735 | 1.03 | 0.1213 |
| LPC O-16_1 | LC | LC-MS database tr,MS1,MS2 | 0.90 | 0.1142 | 1.03 | 0.4160 |
| LPC O-18_0 | LC | LC-MS database tr,MS1,MS2 | 0.95 | 0.1006 | 1.06 | 0.2153 |
| LPC O-18_1 | LC | LC-MS database tr,MS1,MS2 | 0.86 | 0.0835 | 0.96 | 0.2108 |
| LPC P-18_0 | LC | LC-MS database tr,MS1,MS2 | 0.95 | 0.1808 | 0.99 | 0.4250 |
| LPC P-18_1 | LC | LC-MS database tr,MS1,MS2 | 0.91 | 0.1036 | 1.04 | 0.4549 |
| LPE 16_0 | LC | LC-MS database tr,MS1,MS2 | 0.92 | 0.0599 | 0.92 | 0.6848 |

| | | | | | | |
|---|---|---|---|---|---|---|
| LPE 16_1 | LC | LC-MS database tr,MS1,MS2 | 0.97 | 0.9571 | 0.75 | 0.0650 |
| LPE 18_0 sn-1 | LC | LC-MS database tr,MS1,MS2 | 1.03 | 0.5313 | 1.03 | 0.4641 |
| LPE 18_0 sn-2 | LC | LC-MS database tr,MS1,MS2 | 1.04 | 0.9253 | 0.99 | 0.8612 |
| LPE 18_1 sn-1 | LC | LC-MS database tr,MS1,MS2 | 0.94 | 0.2930 | 0.92 | 0.1354 |
| LPE 18_1 sn-2 | LC | LC-MS database tr,MS1,MS2 | 1.07 | 0.9176 | 0.97 | 0.3802 |
| LPE 18_2 sn-1 | LC | LC-MS database tr,MS1,MS2 | 0.93 | 0.8818 | 0.87 | 0.3218 |
| LPE 18_2 sn-2 | LC | LC-MS database tr,MS1,MS2 | 1.05 | 0.9932 | 0.98 | 0.7260 |
| LPE 20_3 | LC | LC-MS database tr,MS1,MS2 | 1.06 | 0.6745 | 1.00 | 0.6251 |
| LPE 20_4 sn-1 | LC | LC-MS database tr,MS1,MS2 | 0.94 | 0.4691 | 0.98 | 0.5219 |
| LPE 20_4 sn-2 | LC | LC-MS database tr,MS1,MS2 | 1.02 | 0.9924 | 1.08 | 0.4245 |
| LPE 20_5 sn-1 | LC | LC-MS database tr,MS1,MS2 | 0.95 | 0.8762 | 0.83 | 0.1769 |
| LPE 20_5 sn-2 | LC | LC-MS database tr,MS1,MS2 | 1.10 | 0.4618 | 0.86 | 0.2471 |
| LPE 22_4 sn-1 | LC | LC-MS database tr,MS1,MS2 | 0.98 | 0.6266 | 0.91 | 0.1505 |
| LPE 22_4 sn-2 | LC | LC-MS database tr,MS1,MS2 | 1.15 | 0.1401 | 1.09 | 0.4243 |
| L-Thyroxine | LC | LC-MS database tr,MS1,MS2 | 1.13 | 0.0959 | 0.94 | 0.0369 |
| Myoinositol | LC | LC-MS database tr,MS1,MS2 | 0.84 | 0.0001 | 1.00 | 0.8660 |
| PC 30_0 | LC | LC-MS database tr,MS1,MS2 | 1.06 | 0.6081 | 0.81 | 0.2736 |
| PC 31_0 | LC | LC-MS database tr,MS1,MS2 | 1.15 | 0.0421 | 0.91 | 0.8110 |
| PC 32_0 | LC | LC-MS database tr,MS1,MS2 | 1.18 | 0.0419 | 0.91 | 0.9913 |
| PC 32_1 | LC | LC-MS database tr,MS1,MS2 | 1.11 | 0.0132 | 1.01 | 0.2837 |
| PC 32_2 | LC | LC-MS database tr,MS1,MS2 | 1.01 | 0.8286 | 0.95 | 0.6154 |
| PC 33_1 | LC | LC-MS database tr,MS1,MS2 | 1.13 | 0.0495 | 1.07 | 0.9304 |
| PC 33_2 | LC | LC-MS database tr,MS1,MS2 | 1.04 | 0.6794 | 0.99 | 0.3082 |
| PC 34_1 | LC | LC-MS database tr,MS1,MS2 | 1.10 | 0.1596 | 1.00 | 0.9043 |
| PC 34_2 | LC | LC-MS database tr,MS1,MS2 | 1.13 | 0.2953 | 1.09 | 0.4972 |
| PC 34_3 | LC | LC-MS database tr,MS1,MS2 | 1.03 | 0.1701 | 0.96 | 0.7896 |
| PC 34_4 | LC | LC-MS database tr,MS1,MS2 | 0.99 | 0.8766 | 1.07 | 0.5969 |
| PC 35_2 | LC | LC-MS database tr,MS1,MS2 | 1.07 | 0.1960 | 1.08 | 0.1905 |
| PC 35_3 | LC | LC-MS database tr,MS1,MS2 | 1.06 | 0.1907 | 1.00 | 0.4779 |
| PC 36_1 | LC | LC-MS database tr,MS1,MS2 | 1.10 | 0.0828 | 0.85 | 0.1119 |
| PC 36_2 | LC | LC-MS database tr,MS1,MS2 | 1.02 | 0.6872 | 1.01 | 0.9453 |
| PC 36_3 | LC | LC-MS database tr,MS1,MS2 | 1.09 | 0.1474 | 1.03 | 0.2709 |
| PC 36_4 | LC | LC-MS database tr,MS1,MS2 | 1.11 | 0.1310 | 1.12 | 0.2647 |
| PC 36_5 | LC | LC-MS database tr,MS1,MS2 | 1.04 | 0.0174 | 0.91 | 0.4970 |
| PC 36_6 | LC | LC-MS database tr,MS1,MS2 | 1.07 | 0.5103 | 1.11 | 0.7955 |

| | | | | | | |
|---|---|---|---|---|---|---|
| PC 38_2 | LC | LC-MS database tr,MS1,MS2 | 1.06 | 0.3560 | 0.85 | 0.3761 |
| PC 38_3 | LC | LC-MS database tr,MS1,MS2 | 1.08 | 0.0507 | 1.00 | 0.9450 |
| PC 38_4 | LC | LC-MS database tr,MS1,MS2 | 1.03 | 0.3320 | 1.06 | 0.6390 |
| PC 38_5 | LC | LC-MS database tr,MS1,MS2 | 1.12 | 0.0268 | 1.06 | 0.1737 |
| PC 38_6 | LC | LC-MS database tr,MS1,MS2 | 1.16 | 0.0028 | 1.11 | 0.2304 |
| PC 38_7 | LC | LC-MS database tr,MS1,MS2 | 1.03 | 0.3708 | 1.06 | 0.6475 |
| PC O-34_2 | LC | LC-MS database tr,MS1,MS2 | 1.06 | 0.4940 | 1.17 | 0.0703 |
| PC O-36_4 | LC | LC-MS database tr,MS1,MS2 | 1.07 | 0.1865 | 1.16 | 0.0690 |
| PC O-36_5 | LC | LC-MS database tr,MS1,MS2 | 1.11 | 0.1277 | 0.97 | 0.8407 |
| PC O-38_5 | LC | LC-MS database tr,MS1,MS2 | 1.09 | 0.3727 | 1.16 | 0.0398 |
| PC O-38_6 | LC | LC-MS database tr,MS1,MS2 | 1.03 | 0.4235 | 1.15 | 0.0747 |
| P-cresol sulfate | LC | standard | 1.15 | 0.0004 | 0.84 | 0.0141 |
| PE 34_2 | LC | LC-MS database tr,MS1,MS2 | 0.99 | 0.3689 | 0.86 | 0.1744 |
| PE 38_6 | LC | LC-MS database tr,MS1,MS2 | 1.12 | 0.0510 | 0.95 | 0.5224 |
| PE O-34_3 | LC | LC-MS database tr,MS1,MS2 | 1.05 | 0.6829 | 1.08 | 0.5806 |
| PE O-36_5 | LC | LC-MS database tr,MS1,MS2 | 0.99 | 0.5336 | 1.03 | 0.7989 |
| PE O-38_6 | LC | LC-MS database tr,MS1,MS2 | 1.10 | 0.2664 | 1.00 | 0.6674 |
| PE O-38_7 | LC | LC-MS database tr,MS1,MS2 | 1.07 | 0.3128 | 1.10 | 0.2890 |
| Phenyl sulfate | LC | standard | 1.16 | 0.0002 | 0.73 | 0.1174 |
| Pyruvic acid | LC | standard | 0.87 | 0.0606 | 0.84 | 0.5128 |
| Resorcinol | LC | LC-MS database tr,MS1,MS2 | 1.10 | 0.8082 | 1.16 | 0.8640 |
| SM 32_1 | LC | LC-MS database tr,MS1,MS2 | 1.16 | 0.0721 | 0.98 | 0.9230 |
| SM 32_2 | LC | LC-MS database tr,MS1,MS2 | 1.09 | 0.0583 | 0.96 | 0.4501 |
| SM 33_1 | LC | LC-MS database tr,MS1,MS2 | 1.25 | 0.0094 | 1.12 | 0.0959 |
| SM 33_2 | LC | LC-MS database tr,MS1,MS2 | 1.13 | 0.0472 | 1.08 | 0.0779 |
| SM 34_0 | LC | standard | 1.15 | 0.1433 | 0.99 | 0.9877 |
| SM 34_1 | LC | LC-MS database tr,MS1,MS2 | 1.17 | 0.0131 | 1.10 | 0.1878 |
| SM 34_2 | LC | LC-MS database tr,MS1,MS2 | 1.09 | 0.1157 | 1.07 | 0.3178 |
| SM 35_2 | LC | LC-MS database tr,MS1,MS2 | 1.13 | 0.1023 | 1.10 | 0.2420 |
| SM 36_1 | LC | LC-MS database tr,MS1,MS2 | 1.23 | 0.0293 | 1.11 | 0.7186 |
| SM 36_2 | LC | LC-MS database tr,MS1,MS2 | 1.14 | 0.3808 | 1.13 | 0.3335 |
| SM 36_3 | LC | LC-MS database tr,MS1,MS2 | 1.09 | 0.0728 | 1.10 | 0.1980 |
| Sphingosine | LC | standard | 0.94 | 0.2220 | 1.03 | 0.7051 |
| Sphingosine-1-phosphate | LC | standard | 1.31 | 0.0401 | 1.00 | 0.7465 |
| TCDCA | LC | standard | 1.30 | 0.1497 | 0.69 | 0.1040 |

| | | | | | | |
|---|---|---|---|---|---|---|
| TDCA | LC | standard | 1.00 | 0.7411 | 0.86 | 0.0118 |
| UDCA | LC | standard | 1.02 | 0.7546 | 1.18 | 0.1527 |
| Uric acid | LC | standard | 1.02 | 0.1100 | 0.98 | 0.1238 |
| Uridine | LC | LC-MS database tr,MS1,MS2 | 1.19 | 0.0004 | 1.08 | 0.1395 |
| Xanthine | LC | standard | 1.87 | 0.0000 | 1.12 | 0.0140 |
| 1-Methylnicotinamide | CE | CE-MS database | 0.91 | 0.6458 | 0.91 | 0.7988 |
| 2-Aminoadipic acid | CE | CE-MS database | 1.36 | 0.0050 | 0.95 | 0.9113 |
| 2-Aminobutyric acid | CE | CE-MS database | 0.97 | 0.5904 | 1.16 | 0.0890 |
| 2-Hydroxy-4-methylvaleric acid | CE | CE-MS database | 1.57 | 0.0001 | 0.99 | 0.6309 |
| 2-Hydroxybutyric acid | CE | CE-MS database | 1.37 | 0.0000 | 1.14 | 0.1207 |
| 2-Hydroxyvaleric acid | CE | CE-MS database | 1.16 | 0.0006 | 1.13 | 0.0843 |
| 2-Oxovaleric acid | CE | CE-MS database | 1.69 | 0.0000 | 1.19 | 0.5942 |
| 3 or 4-Methyl-2-oxovaleric acid | CE | CE-MS database | 2.00 | 0.0000 | 1.12 | 0.0345 |
| 3-Hydroxybutyric acid | CE | CE-MS database | 1.28 | 0.0718 | 0.93 | 0.7302 |
| 3-Indoxylsulfuric acid | CE | CE-MS database | 1.39 | 0.0000 | 0.91 | 0.1184 |
| 3-Methylhistidine | CE | CE-MS database | 1.04 | 0.1219 | 0.81 | 0.7722 |
| 3-Phenylpropionic acid | CE | CE-MS database | 1.00 | 0.5905 | 1.00 | 0.5022 |
| 4-Guanidinobutyric acid | CE | CE-MS database | 1.13 | 0.4381 | 0.96 | 0.2290 |
| 5-Methoxyindoleacetic acid | CE | CE-MS database | 1.44 | 0.0018 | 1.20 | 0.1788 |
| 5-Oxoproline | CE | CE-MS database | 1.01 | 0.8972 | 0.92 | 0.8717 |
| ADMA | CE | CE-MS database | 1.10 | 0.5413 | 1.03 | 0.0818 |
| ADP | CE | CE-MS database | 1.95 | 0.0088 | 0.93 | 0.3954 |
| Ala | CE | CE-MS database | 1.42 | 0.0000 | 1.04 | 0.6977 |
| AMP | CE | CE-MS database | 1.96 | 0.0093 | 0.67 | 0.3310 |
| Arg | CE | CE-MS database | 1.23 | 0.0098 | 1.01 | 0.4822 |
| Asn | CE | CE-MS database | 1.04 | 0.2971 | 1.09 | 0.6355 |
| Asp | CE | CE-MS database | 1.25 | 0.0497 | 1.02 | 0.7508 |
| Benzoic acid | CE | CE-MS database | 1.19 | 0.2076 | 1.10 | 0.5963 |
| Betaine | CE | CE-MS database | 0.74 | 0.3921 | 0.90 | 0.8407 |
| Butyric acid | CE | CE-MS database | 1.19 | 0.0020 | 1.05 | 0.2497 |
| Carboxymethyllysine | CE | CE-MS database | 1.08 | 0.7078 | 1.10 | 0.2275 |
| Citric acid | CE | CE-MS database | 1.26 | 0.0000 | 1.02 | 0.7016 |
| Citrulline | CE | CE-MS database | 1.03 | 0.2906 | 0.98 | 0.7529 |
| Creatine | CE | CE-MS database | 1.50 | 0.0001 | 0.95 | 0.9100 |
| Ethanolamine phosphate | CE | CE-MS database | 1.18 | 0.3486 | 0.86 | 0.6423 |

| | | | | | | |
|---|---|---|---|---|---|---|
| Gln | CE | CE-MS database | 0.98 | 0.8423 | 1.02 | 0.3816 |
| Glu | CE | CE-MS database | 1.37 | 0.0197 | 0.88 | 0.4354 |
| Gluconic acid | CE | CE-MS database | 0.96 | 0.0198 | 1.04 | 0.6920 |
| Glucosamine | CE | CE-MS database | 1.40 | 0.5028 | 1.30 | 0.9896 |
| Gly | CE | CE-MS database | 1.04 | 0.3547 | 0.98 | 0.3761 |
| Glyceric acid | CE | CE-MS database | 0.91 | 0.0104 | 1.04 | 0.8819 |
| Glycerol 3-phosphate | CE | CE-MS database | 1.30 | 0.0004 | 0.92 | 0.3958 |
| Glycerophosphocholine | CE | CE-MS database | 1.22 | 0.2309 | 0.75 | 0.3844 |
| Glyoxylic acid | CE | CE-MS database | 0.97 | 0.5130 | 1.07 | 0.0356 |
| Guanidoacetic acid | CE | CE-MS database | 1.33 | 0.0469 | 1.02 | 0.3278 |
| Heptanoic acid | CE | CE-MS database | 1.15 | 0.5082 | 0.94 | 0.4158 |
| Hexanoic acid | CE | CE-MS database | 1.11 | 0.0631 | 0.95 | 0.9270 |
| His | CE | CE-MS database | 1.63 | 0.0045 | 0.94 | 0.3638 |
| Homoarginine | CE | CE-MS database | 1.16 | 0.0873 | 0.90 | 0.9532 |
| Hydroxyproline | CE | CE-MS database | 1.03 | 0.2574 | 0.97 | 0.5691 |
| Hypoxanthine | CE | CE-MS database | 4.39 | 0.0000 | 0.80 | 0.4181 |
| Ile | CE | CE-MS database | 1.09 | 0.1509 | 0.95 | 0.2438 |
| Indole-3-acetic acid | CE | CE-MS database | 1.04 | 0.2150 | 0.91 | 0.6582 |
| Isocitric acid | CE | CE-MS database | 1.61 | 0.0000 | 1.05 | 0.5887 |
| Isovaleric acid | CE | CE-MS database | 1.23 | 0.0041 | 0.90 | 0.4647 |
| Kynurenine | CE | CE-MS database | 1.04 | 0.6543 | 1.07 | 0.3746 |
| Lactic acid | CE | CE-MS database | 4.06 | 0.0000 | 1.03 | 0.8139 |
| Leu | CE | CE-MS database | 1.13 | 0.1831 | 1.03 | 0.3552 |
| Lys | CE | CE-MS database | 1.37 | 0.0102 | 0.99 | 0.5136 |
| Malic acid | CE | CE-MS database | 1.71 | 0.0000 | 1.08 | 0.1538 |
| Met | CE | CE-MS database | 1.79 | 0.0000 | 1.39 | 0.1614 |
| Methionine sulfoxide | CE | CE-MS database | 1.05 | 0.4762 | 1.06 | 0.4830 |
| Mucic acid | CE | CE-MS database | 1.02 | 0.5709 | 0.84 | 0.2458 |
| N,N-Dimethylglycine | CE | CE-MS database | 1.08 | 0.8579 | 0.98 | 0.2718 |
| N2-Phenylacetylglutamine | CE | CE-MS database | 1.35 | 0.0001 | 0.85 | 0.0772 |
| N5-Ethylglutamine | CE | CE-MS database | 1.14 | 0.4313 | 1.59 | 0.0553 |
| N6,N6,N6-Trimethyllysine | CE | CE-MS database | 0.90 | 0.8617 | 0.83 | 0.1802 |
| N6-Methyllysine | CE | CE-MS database | 1.04 | 0.1195 | 1.13 | 0.3247 |
| N-Acetylalanine | CE | CE-MS database | 1.24 | 0.0264 | 1.02 | 0.2144 |
| N-Acetylglycine | CE | CE-MS database | 1.00 | 0.1458 | 1.18 | 0.1832 |

| | | | | | | |
|---|---|---|---|---|---|---|
| N-Acetylmuramic acid | CE | CE-MS database | 1.24 | 0.0509 | 1.10 | 0.7505 |
| N-Methylproline | CE | CE-MS database | 1.02 | 0.9238 | 0.58 | 0.4894 |
| Octanoic acid | CE | CE-MS database | 1.54 | 0.0001 | 0.97 | 0.7812 |
| Ornithine | CE | CE-MS database | 1.23 | 0.0291 | 0.94 | 0.7299 |
| Pantothenic acid | CE | CE-MS database | 1.38 | 0.4434 | 0.85 | 0.0207 |
| Phe | CE | CE-MS database | 1.17 | 0.0607 | 1.05 | 0.2509 |
| Pipecolic acid | CE | CE-MS database | 1.07 | 0.3789 | 1.15 | 0.3144 |
| Pro | CE | CE-MS database | 1.42 | 0.0000 | 1.00 | 0.8703 |
| Pyrophosphate | CE | CE-MS database | 1.12 | 0.8980 | 1.03 | 0.6989 |
| SDMA | CE | CE-MS database | 1.16 | 0.0228 | 1.16 | 0.0409 |
| Ser | CE | CE-MS database | 1.02 | 0.3987 | 1.05 | 0.2843 |
| S-Methylcysteine | CE | CE-MS database | 1.43 | 0.0056 | 0.99 | 0.3515 |
| Stachydrine | CE | CE-MS database | 0.90 | 0.2607 | 0.58 | 0.6833 |
| Succinic acid | CE | CE-MS database | 1.69 | 0.0000 | 0.98 | 0.6927 |
| Taurine | CE | CE-MS database | 1.16 | 0.0704 | 1.07 | 0.9713 |
| Thr | CE | CE-MS database | 0.99 | 0.5725 | 1.02 | 0.5005 |
| Threonic acid | CE | CE-MS database | 0.93 | 0.5496 | 0.91 | 0.2408 |
| Trigonelline | CE | CE-MS database | 0.96 | 0.9366 | 1.73 | 0.1381 |
| Trp | CE | CE-MS database | 1.07 | 0.2023 | 1.03 | 0.3844 |
| Tyr | CE | CE-MS database | 1.10 | 0.0942 | 1.00 | 0.5238 |
| Urea | CE | CE-MS database | 1.01 | 0.3904 | 1.01 | 0.9974 |
| Val | CE | CE-MS database | 1.05 | 0.1172 | 1.00 | 0.4286 |
| γ-Butyrobetaine | CE | CE-MS database | 1.14 | 0.0114 | 1.11 | 0.5947 |

*Metabolites were identified using either standards added before sample extraction, or a CE-MS database containing 960 metabolite standards provided by Human Metabolome Technologies, Inc. (HMT), or an in-house LC-MS database with retention time (tr), MS1 and MS2.*

ESM Figure

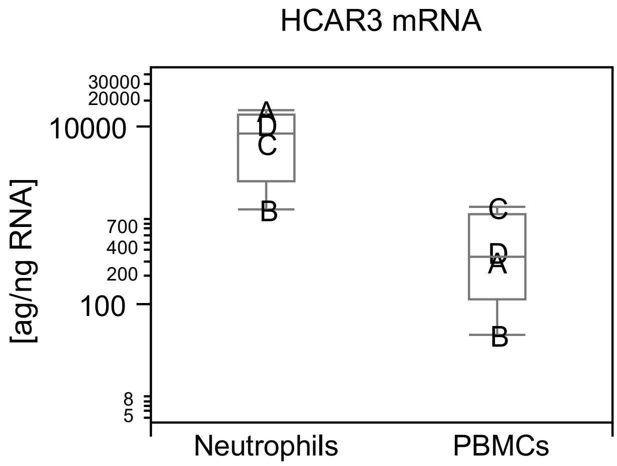


**Fig. 1:** Transcript levels of the Hydroxycarboxylic acid receptor 3 (HCAR3) in peripheral blood mononuclear cells (PBMCs) and granulocytes. Letters denote different donors. HCAR3 levels in individual samples were quantified using a standard with known concentration and expressed relative to total RT-PCR input RNA.
